# Long-term air pollution exposure and markers of cardiometabolic health in the National Longitudinal Study of Adolescent to Adult Health (Add Health) Study

**DOI:** 10.1101/2022.12.07.22283112

**Authors:** Mercedes A. Bravo, Fang Fang, Dana B. Hancock, Eric O. Johnson, Kathleen Mullan Harris

## Abstract

**Background:** Air pollution exposure is associated with cardiovascular morbidity and mortality. Although exposure to air pollution early in life may represent a critical window for development of cardiovascular disease risk factors, few studies have examined associations of long-term air pollution exposure with markers of cardiovascular and cardiometabolic health in young adults.

**Objectives:** By combining health data from the National Longitudinal Study of Adolescent to Adult Health (Add Health) with air pollution data from the Fused Air Quality Surface using Downscaling (FAQSD) archive, we: (1) calculated multi-year estimates of exposure to ozone (O_3_) and particulate matter with an aerodynamic diameter ≤2.5μm (PM_2.5_) for Add Health participants; and (2) estimated associations between air pollution exposures and multiple markers of cardiometabolic health.

**Methods:** Add Health is a nationally representative longitudinal cohort study of over 20,000 adolescents aged 12–19 in the United States (US) in 1994–95 (Wave I). Participants have been followed through adolescence and into adulthood with five in-home interviews. Estimated daily concentrations of O_3_ and PM_2.5_ at census tracts were obtained from the FAQSD archive and used to generate tract-level annual averages of O_3_ and PM_2.5_ concentrations. We estimated associations between average O_3_ and PM_2.5_ exposures from 2002–07 and markers of cardiometabolic health measured at Wave IV (2008–09), including hypertension, hyperlipidemia, body mass index (BMI), diabetes, C-reactive protein, and metabolic syndrome.

**Results:** The final sample size was 11,259 individual participants. The average age of participants at Wave IV was 28.4 years (range: 24–34 years). In models adjusting for age, race/ethnicity, and sex, long-term O_3_ exposure (2002–07) was associated with elevated odds of hypertension, with an odds ratio (OR) of 1.015 (95% confidence interval [CI]: 1.011, 1.029); obesity (1.022 [1.004, 1.040]); diabetes (1.032 [1.009,1.054]); and metabolic syndrome (1.028 [1.014, 1.041]); PM_2.5_ exposure (2002–07) was associated with elevated odds of hypertension (1.022 [1.001, 1.045]).

**Conclusion:** Findings suggest that long-term ambient air pollution exposure, particularly O_3_ exposure, is associated with cardiometabolic health in early adulthood.

## INTRODUCTION

An extensive literature has demonstrated that exposure to ambient air pollution has wide-ranging harmful effects on health.^1^ Exposures to ozone (O_3_) and particulate matter with an aerodynamic diameter ≤2.5μm (PM_2.5_) are linked with cardiovascular and respiratory-related mortality,^2-4^ morbidity, ^5-8^ and hospital admissions,^9,10^ even at concentrations below the National Ambient Air Quality Standards.^11,12^ A growing body of research has investigated the relationship between air pollution exposure, particularly traffic-related air pollution exposure, and risk factors for early markers of cardiovascular disease. Evidence increasingly suggests that exposure to air pollutants is associated with risk factors for early indicators of cardiovascular disease that may develop years or decades prior to clinical manifestations of more severe disease.^13^

Specifically, chronic exposure to air pollution early in the life course may directly affect development of major risk factors for cardiovascular disease, including obesity, hypertension, and metabolic disorders. Studies have documented associations between residential proximity to roads and arterial stiffness,^14^ an early-stage indicator of atherosclerosis, and the underlying cause of most cardiovascular disease. There is also evidence to suggest that long-term exposure to air pollution, including PM_2.5_ and O_3_, is associated with accelerated atherosclerosis,^15^ elevated blood pressure and increased mean arterial pressure,^16^, obesity,^17,18^ and diabetes.^19,20^ Much of the work on air pollution and cardiovascular disease risk has focused on middle-aged and older adults, e.g., ≥45 years, the demographic age group at highest risk for morbidity and mortality associated with cardiovascular disease.

However, exposure to air pollution during early developmental stages (childhood, adolescence, and early adulthood) may represent critical windows for development of risk factors for cardiovascular disease. For example, a 2021 study of children in Los Angeles, CA, observed that children with higher exposure to traffic-related air pollution had greater annual change in carotid artery intima-media thickness, a measure indicative of subclinical atherosclerosis.^21^ Jerrett et al. examined traffic pollution around children’s residences and observed that higher levels of traffic-related air pollution were associated with higher body mass index (BMI) in children 10-18 years, particularly in females.^22^ Li et al. studied adults aged 18 to 29 and found that high concentrations of some air pollutants were associated with impairments in high-density lipoprotein (HDL) functionality,^23^ and McGuinn et al. found that long-term PM_2.5_ exposure was associated with lipoprotein increases in adults.^24^ Studies of air pollution exposure and cardiometabolic outcomes in children, adolescents, or young adults are few, often have shorter follow up periods, and tend to be based on small or non-representative samples in localized geographic areas.^23,25,26^

The National Longitudinal Study of Adolescent to Adult Health (Add Health) is a nationally representative cohort of 20,745 individuals in the United States (US). Add Health participants were enrolled as adolescents aged 12-19 years in 1994–95 and have been followed through adolescence and into adulthood until, most recently, 2016–18.^27^ Add Health and its collection of individual-level measures of cardiometabolic health offer an important and unique opportunity to further our understanding of potential adverse health effects of air pollution exposures that occur in early adulthood. Markers of cardiometabolic health can provide insight into underlying biological processes in premorbid disease pathways and may indicate biological dysfunction before clinical manifestation of more severe disease.

In this study, we used publicly available daily estimates of ambient concentrations of two criteria air pollutants, PM_2.5_ and O_3_, to generate annual average exposure estimates, and merged these to Add Health participants based on census tract(s) of residence across the multiple waves of Add Health. We then: (1) evaluated whether multi-year averages of air pollutant exposure prior to Wave IV, which occurred in 2008–09, differed by individual-level characteristics (e.g., race/ethnicity, sex); and (2) estimated associations between air pollution exposure and six markers of cardiometabolic health measured at Wave IV: hypertension, hyperlipidemia, obesity, diabetes, C-reactive protein, and a summary measure of metabolic syndrome. This is the first study of long-term air pollution exposure in Add Health and, to the best of our knowledge, one of a limited number of studies examining air pollution exposures and cardiometabolic health in a cohort of young adults.^26^

## METHODS

### Add Health cohort

We used data from the National Longitudinal Study of Adolescent to Adult Health (Add Health), an ongoing, nationally representative cohort of adolescents in grades 7–12 during the 1994–95 school year. Add Health used a multistage, stratified, school-based, cluster sampling design to select a probability sample of more than 20,000 adolescents from school rosters for in-home interviews at Wave I (WI) in 1994–95 (ages 12–19). The cohort has been followed up with four subsequent interviews: WII in 1996 (ages 13–20); WIII in 2001–02 (ages 18–26); WIV in 2008–09 (ages 24–32); and WV in 2016–18 (ages 33–43).^27,28^ Response rates have ranged from 72% to 90% across waves, and non-response bias has been minimal.^29,30^ Details on the Add Health study objectives, design, and sampling strategy are provided elsewhere.^27,28^

For the exposure measures, the present paper uses Add Health participant data from Waves I, III, and IV with recently constructed and merged air pollution data, as described later. For the cardiometabolic health markers, we used the biological and clinical data collected at Wave IV (response rate=80.3%), including systolic and diastolic blood pressure; lipid panels; measured height and weight used to calculate BMI; indicators of diabetes (e.g., fasting or non-fasting glucose and/or glycated hemoglobin [HbA1c]); and concentrations of C-reactive protein as a measure of inflammation.^31^ We examined six key markers of cardiometabolic health at Wave IV: hypertension, hyperlipidemia, obesity, diabetes, and C-reactive protein concentration, and a summary measure of metabolic syndrome. Following clinical guidelines to identify the high-risk category, all cardiometabolic health markers were coded as binary variables (0/1) and defined as follows.

Hypertension was indicated if participants met at least one of the following criteria: having stage 1 or stage 2 hypertension according to the guidelines from the Seventh Report of the Joint National Committee on Prevention Detection, Evaluation, and Treatment of High Blood Pressure (JNC7): systolic blood pressure of 140–159 mm Hg or diastolic blood pressure of 80–89 mm Hg for stage 1 hypertension; systolic blood pressure of ≥160 mm Hg or diastolic blood pressure of ≥100 mm Hg for stage 2 hypertension;^32^ or self-reported history of having physician-diagnosed high blood pressure; or self-reported anti-hypertensive medication use.

Hyperlipidemia was indicated if participants had a self-reported history of physician-diagnosed high cholesterol or triglycerides; or self-reported anti-hyperlipidemic medication use in the past 4 weeks.

Participants’ BMI was calculated based on height and weight measured at Wave IV as kg/m^2^. Respondents were considered obese if BMI was ≥30.

Diabetes was indicated if participants met at least one of the following criteria: either fasting glucose ≥126 mg/dL or non-fasting glucose ≥200 mg/dL or glycated hemoglobin (HbA1c) ≥6.5%; or self-reported history of having physician-diagnosed diabetes except during pregnancy; or self-reported anti-diabetic medication use.

Inflammation was measured by high-sensitivity C-reactive protein (hsCRP) and coded as a binary indicator of high inflammation if hsCRP was > 3mg/L. C-reactive protein is a widely measured biomarker of systemic inflammation, a well-established risk factor for the development and progression of cardiometabolic diseases.^33,34^

Metabolic syndrome represents a cluster of conditions that increases the risk of heart disease, stroke, and diabetes.^35,36^ Metabolic syndrome was indicated if participants had three or more of the following conditions: (1) blood pressure >130/85 mm Hg, or self-reported physician-diagnosed hypertension, or self-reported anti-hypertensive medication usage; (2) HbA1c ≥ 5.7%; (3) membership in the lowest two high-density lipoprotein (HDL) cholesterol deciles for women or the lowest three HDL deciles for men; (4) membership in the top three deciles of triglycerides; and (5) waist circumference ≥88cm for women or ≥102cm for men. This definition of metabolic syndrome has been used in previous studies of Add Health participants.^37^

### Analytic Samples/Inclusion Criteria

We created an analytical dataset from the Add Health study sample at Wave I (n=20,745). We included only Wave I individuals who participated in Wave III *and* Wave IV; were geocoded; resided in the continental US (individuals residing in Alaska or Hawaii were excluded because air pollution data was not available for these states); and had non-missing data on key covariates, i.e., age, race/ethnicity, and sex (Figure 1). We also included only US-born participants, due to health differences by nativity status,^38-41^ and women who were not pregnant at Wave IV, as anthropometric and physiological markers differ for pregnant vs. non-pregnant women.

**Figure 1.**
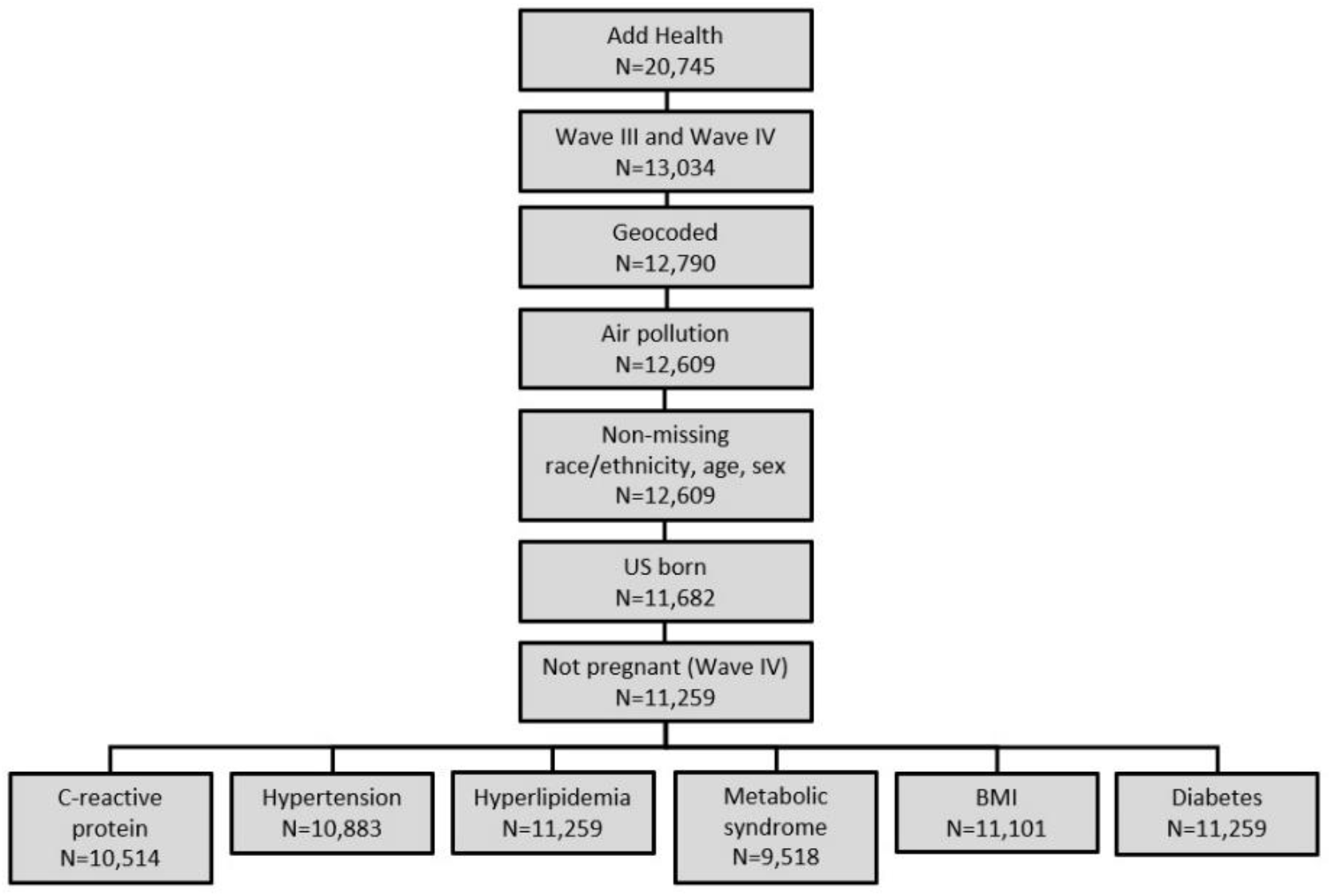
Sample size restrictions flow chart.

With these inclusion criteria, a maximum of 11,259 participants remained for statistical analyses. Sample sizes varied slightly across analyses depending on the health outcome, due to missing health measurements for some markers of cardiometabolic health (Figure 1).

### Air pollution exposure

Air pollution exposure estimates were generated using the publicly available Fused Air Quality Surface using Downscaling (FAQSD) files (https://www.epa.gov/hesc/rsig-related-downloadable-data-files). The FAQSD files include daily predictions of 24-hour average PM_2.5_ concentrations and 8-hour maximum O_3_ concentrations at 2010 US Census tract centroids. The FAQSD files were generated using a statistical model (“downscaler”) that relates monitoring data and gridded output from the Community Multiscale Air Quality (CMAQ) model using a linear regression model with additive and multiplicative bias coefficients that can vary in space and time (see Supplemental Information for additional details).^42,43^

FAQSD files included predictions of daily 24-hour average PM_2.5_ concentrations and 8-hour maximum O_3_ concentrations at census tracts for 2002–07. We averaged daily values to generate annual average concentration estimates of tract-level 8-hour maximum O_3_ and 24-hour average PM_2.5_ for each year. Add Health participants were then matched to air pollution values based on their census tract(s) of residence and year. Residential location information on participants was available at each wave, but not for between-wave years, while air pollution data were obtained for 2002–07 (Figure 2). As illustrated in Figure 2, annual average PM_2.5_ and O_3_ concentrations from 2002–05 were attached to Add Health participants based on their census tract of residence at Wave III (2001–02) and annual average PM_2.5_ and O_3_ concentrations from 2006–09 were attached to Add Health participants based on their census tract of residence at Wave IV (2008–09). Because we did not have information on when individuals moved in between-wave years, for those who changed residences we assumed they moved close to the “midpoint” of years between waves.^44^ Thus, we assumed that Add Health participants resided at their Wave III address between 2002–05, and at their Wave IV address between 2006–09.

**Figure 2.**
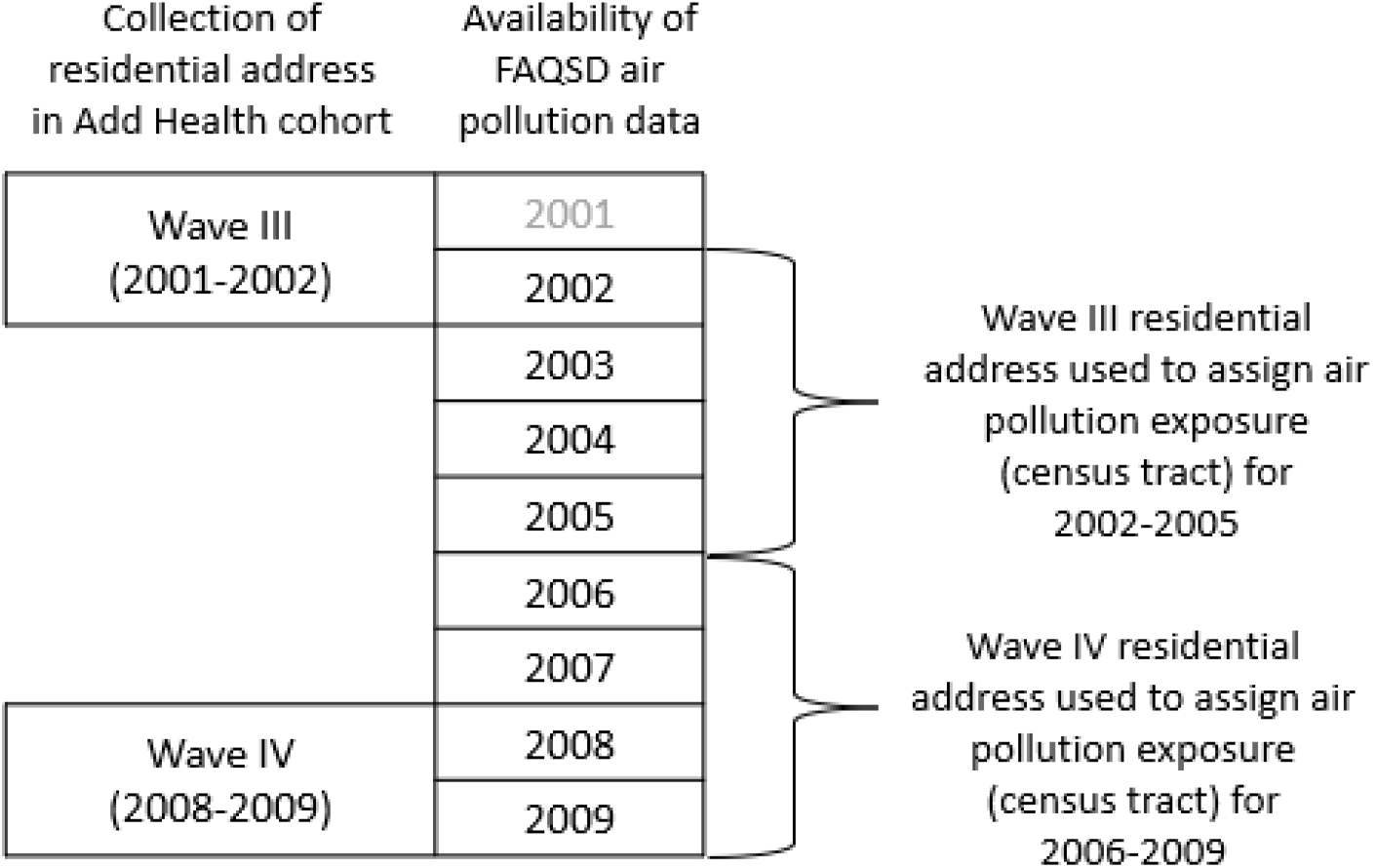
Residential address collection, availability of air pollution data, and assignment of air pollution data to Add Health cohort members^a^. ^a^ FAQSD air pollution data were not available in 2001, which is shown in gray. Air pollution data from 2008 and 2009 were merged with Add Health participant data as shown here, but air pollution data from 2008-2009 were not used in the present study.

We examined air pollution exposure and the key markers of cardiometabolic health at Wave IV in the analytical sample of 11,259 individuals. We used FAQSD output to estimate average PM_2.5_ and O_3_ exposures from 2002–07, which represents the long-term average air pollution exposure prior to collection of cardiometabolic health measures at Wave IV in 2008–09. Because FAQSD data are not available prior to 2002, we are unable to estimate air pollution exposure prior to 2002 using this particular data source.

Written informed consent was obtained at all waves and protocols approved by the Institutional Review Board of the University of North Carolina at Chapel Hill.

### Statistical analysis

We calculated descriptive statistics of the study population, as well as average air pollution exposures by individual-level characteristics (e.g., age, sex, race/ethnicity). We tested for differences in 2002–07 O_3_ and PM_2.5_ concentrations by sex and racial/ethnic group, applying post-hoc tests for multiple pairwise comparisons and adjusting p-values for multiple comparisons using Bonferoni.^45,46^ To examine associations between air pollution exposures and markers of cardiometabolic health among young adults, we fit logistic regression generalized estimating equations (GEEs) to estimate the association between each air pollutant (O_3_, PM_2.5_) and health outcome. Models adjusted for Add Health participant’s age (years), sex, and race/ethnicity (non-Hispanic Black, Non-Hispanic White, Hispanic, Other):

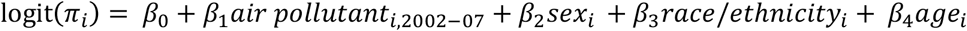

where *π*_*i*_ is the probability of experiencing the outcome of interest for subject *i*. The coefficient *β*_1_ represents the estimated association between the health outcome and air pollution exposure averaged over 2002–07; the coefficients *β*_2,_ *β*_3,_ and *β*_4_ represent the associations of sex, race/ethnicity, and Wave IV age, respectively, with the outcome of interest. Models were fit separately for each pollutant and health outcome. An exchangeable correlation matrix was specified on the primary sampling unit ID in Add Health.^47^ We fit unweighted multivariable models with covariates adjusting for unequal selection into the sample and accounting for the clustered school design using GEEs.^48^

A two-sided α=0.05 was used to signify statistical significance. Statistical analyses were conducted using R statistical software.^49^ The *gee* package was used for fitting GEE models.^50^

### Sensitivity analysis

As a sensitivity analysis, we fit adjusted models that estimated associations between air pollution exposure in 2006–07 and markers of cardiometabolic health measured at Wave IV. Analysis of 2002–07 air pollution exposure requires that individuals participated in both Waves III and IV, because their residential address at each wave is required to assign air pollution exposure. In contrast, analysis of 2006– 07 air pollution exposure only requires that individuals participated in Wave IV, because only a Wave IV address is used to assign air pollution exposure estimates for 2006 and 2007 (e.g., Figure 2). The 2006–07 exposure period represents lagged exposure in the years just prior to Wave IV. We conducted this sensitivity analysis to explore whether results were consistent: (1) across exposure averaging intervals, e.g., 2002–07 and 2006–07; and (2) using the slightly larger sample that is available for the 2006–07 analysis. We used the same set of restriction criteria previously applied to the sample of Wave III and IV participants, resulting in an analytical sample size of n=13,867 individuals for the sensitivity analysis (SM, Figure S1).

## RESULTS

### Descriptive statistics

Descriptive statistics of the study sample are provided in Table 1. At Wave IV, Add Health participants were, on average, 28 years old, and just over half the sample was female. Non-Hispanic White participants comprised over half of the sample (∼66%). The most common cardiometabolic health outcomes were high inflammation and obesity, followed by hypertension and metabolic syndrome.

**Table 1.** Distributions of participants’ characteristics for those who participated in Wave III and Wave IV (n=11,259)^a^.

|  |  |
| --- | --- |
| **Variable** | **Mean value (SD)** |
| Age at exam, Wave IV, years | 28.4 (1.78) |
| O<sub>3</sub> exposure (ppb), 2002-2007 | 38.3 (3.67) |
| PM<sub>2.5</sub> exposure (µg/m<sup>3</sup>), 2002-2007 | 12.7 (2.47) |
|  | **N (%)** |
| Women, % | 5,961 (52.9) |
| Race/ethnicity |  |
| Non-Hispanic Black | 2,550 (22.6) |
| Non-Hispanic White | 6,881 (61.1) |
| Hispanic | 1,281 (11.4) |
| Other | 547 (4.88) |
| Health outcomes |  |
| Hypertension | 2,840 (26.1) |
| Hyperlipidemia | 921 (8.18) |
| Obese (BMI ≥ 30) | 4,192 (37.8) |
| Diabetes | 811 (7.20) |
| High inflammation | 4,074 (38.7) |
| Metabolic syndrome | 2,331 (20.7) |
| <sup>a</sup>Summary statistics are presented for the analytic sample size of Wave III and Wave IV participants (n=11,259). |  |

### Air pollution exposure

Air pollution exposure estimates are summarized in Table 2 overall, and by sex and race/ethnicity. Average O_3_ and PM_2.5_ exposures were 38.3 ppb and 12.7 μg/m^3^, respectively, over the 2002–07 period. T-tests with unequal variances were used to test for differences in O_3_ and PM_2.5_ concentrations by sex and the Kruskal-Wallis test was used to evaluate differences in 2002–07 air pollutant concentrations by racial/ethnic group.^51^ Air pollutant concentrations did not differ by sex, but there were differences between in PM_2.5_ and O_3_ by racial/ethnic group (p<0.0001).

**Table 2.** Distributions of O_3_ and PM_2.5_ exposure by demographic characteristics^a^.

|  |  |  |  |  |
| --- | --- | --- | --- | --- |
|  | **2002-2007**<br>**(n=11,259)** |  |  |  |
|  | **Ozone** |  | **PM<sub>2.5</sub>** |  |
|  | **Concentration (ppb)** | **p-value** | **Concentration (µg/m<sup>3</sup>)** | **p-value** |
| Overall | 38.3 (3.67) |  | 12.7 (2.47) |  |
| Sex |  | 0.426 |  | 0.982 |
| Female | 38.3 (3.67) |  | 12.7 (2.49) |  |
| Male | 38.3 (3.67) |  | 12.7 (2.49) |  |
| Race/ethnicity |  | < 0.0001 |  | < 0.0001 |
| Non-Hispanic Black | 38.6 (3.35) |  | 13.6 (2.05) |  |
| Non-Hispanic White | 38.6 (3.45) |  | 12.2 (2.27) |  |
| Hispanic | 37.0 (4.18) |  | 12.8 (3.28) |  |
| Other | 36.1 (4.85) |  | 13.4 (3.06) |  |
| <sup>a</sup>t-tests with unequal variances were used to test for differences in O<sub>3</sub> and PM<sub>2.5</sub> exposure by sex. The Kruskal Wallis test was used to test for differences in O<sub>3</sub> and PM<sub>2.5</sub> by racial/ethnic group. P-values in the table correspond to those obtained from t-tests (sex) and Kruskal Wallis tests (race/ethnicity). Additionally, a Dunn's test was used for pairwise comparisons of air pollutant concentrations for different racial/ethnic groups. |  |  |  |  |

Non-Hispanic Black and non-Hispanic White groups had the highest O_3_ exposure (38.6 ppb); Other racial/ethnic groups (e.g., Asian, Native American, “other” race) had the lowest O_3_ exposure (36.1 ppb). For O_3_ exposure, Dunn’s test indicated that the non-Hispanic White group had different O_3_ exposures compared to Hispanic (p < 0.0001) and Other groups (p < 0.0001). The non-Hispanic Black group also had different O_3_ exposures compared to Hispanic (p < 0.0001) and Other groups (p < 0.0001). With respect to PM_2.5_, the non-Hispanic Black group had the highest PM_2.5_ exposure (13.6 μg/m^3^), while the non-Hispanic White group had the lowest PM_2.5_ exposure (12.2 μg/m^3^). Dunn’s tests indicated that the non-Hispanic Black group had different PM_2.5_ exposures compared to non-Hispanic White (p < 0.0001) and Hispanic groups (p < 0.0001). The non-Hispanic White group had different PM_2.5_ exposures compared to Hispanic (p < 0.0001) and Other groups (p < 0.0001). PM_2.5_ exposure in the Hispanic group differed from that in the Other group as well (p=0.00112).

### Associations of air pollution exposure and health outcomes

Odds ratios (ORs) for associations of each air pollutant (O_3_ and PM_2.5_) with each health outcome are presented in Table 3. In GEEs adjusting for age, race/ethnicity, and sex, 2002–07 O_3_ exposure was associated with elevated odds of hypertension, with an OR of 1.015 (95% confidence interval [CI]: 1.011, 1.034); obesity (1.022 [1.004, 1.040]); diabetes (1.032 [1.009,1.054]); and metabolic syndrome (1.028 [1.014, 1.041]) (Table 3). In models adjusting for age, race/ethnicity, and sex, 2002–07 PM_2.5_ exposure was associated with elevated odds of hypertension (1.022 [1.001, 1.045]).

**Table 3.** Wave IV health outcomes and air pollution exposure (2002-2007) in adjusted GEE logistic regression models ^a^.

|  |  |  |  |  |
| --- | --- | --- | --- | --- |
|  | **Ozone** |  | **PM<sub>2.5</sub>** |  |
| Health outcome (n) | OR<br>(95% CI) | p-value | OR<br>(95% CI) | p-value |
| Hypertension (n=10,883) | **1.015**<br>**(1.011, 1.029)** | **0.0342** | **1.022**<br>**(1.001, 1.045)** | **0.0457** |
| Hyperlipidemia (n=11,259) | 1.009<br>(0.991, 1.028) | 0.310 | 1.013<br>(0.986, 1.041) | 0.353 |
| Obese (n=11,101) | **1.022**<br>**(1.004, 1.040)** | **0.0192** | 0.991<br>(0.965, 1.017) | 0.476 |
| Diabetes (n=11,259) | **1.032**<br>**(1.009, 1.054)** | **0.00502** | 0.975<br>(0.938, 1.014) | 0.208 |
| Inflammation (n=10,514) | 1.012<br>(0.999, 1.025) | 0.0627 | 1.003<br>(0.982, 1.025) | 0.759 |
| Metabolic syndrome (n=9,518) | **1.028**<br>**(1.014, 1.041)** | **0.0000770** | 0.994<br>(0.967, 1.022) | 0.692 |
| <sup>a</sup>GEEs were fit as logistic regression models with a binary health outcome, Primary Sampling Unit School Identifier (PSUSCID) as the cluster ID variable, and an exchangeable correlation matrix. Models adjusted for 2002-07 pollutant concentration, age at Wave IV (years), sex (female or male), and race/ethnicity (non-Hispanic Black, non-Hispanic White, Hispanic, and Other). |  |  |  |  |

### Sensitivity analysis

Summary statistics of the analytic sample examined for 2006–07 exposure are provided in SM Table S1. Exposure to O_3_ in 2006–07 was associated with elevated odds of obesity (1.014 [1.003, 1.025]); diabetes (1.024 [1.004, 1.045]); inflammation (1.012 [1.003, 1.022]); and metabolic syndrome (1.013 [1.001, 1.026]) (SM Table S2). Statistically significant associations were not observed between 2006–07 PM_2.5_ exposure and health outcomes, although there was a marginal association of PM_2.5_ with hypertension (p=0.0518).

## DISCUSSION

Add Health is a longstanding, nationally representative cohort of individuals with rich longitudinal data and a follow up period of nearly 25 years. Here, for the first time, we leverage the wealth of data available in this cohort to evaluate health outcomes in early adulthood that may be associated with long-term exposure to air pollution. Specifically, we: (1) examined whether multi-year exposures to O_3_ and PM_2.5_ differed by individual-level characteristics; and (2) estimated associations between air pollution exposure during the cohort’s early to mid-20s and multiple markers of cardiometabolic health measured when the cohort entered their late 20s.

We observed associations of 2002–07 O_3_ exposure with elevated odds of hypertension, obesity, diabetes, and metabolic syndrome. Results were similar for a shorter 2-year lagged period (2006–07) O_3_ exposure, which was associated with elevated odds of obesity, diabetes, metabolic syndrome, and inflammation. Long-term exposure to PM_2.5_ (2002–07) was associated with elevated odds of hypertension, and no statistically significant associations were observed between 2006 –07 PM_2.5_ exposure and health outcomes examined in our study.

Associations were in the expected direction, i.e., higher air pollution levels were associated with elevated odds of cardiometabolic outcomes, and we observed stronger evidence for associations between health outcomes and O_3_ compared to PM_2.5_.^52^ This is notable because the current body of evidence for cardiovascular and cardiometabolic health outcomes is strongest for PM_2.5_ exposure, although associations between O_3_ and cardiometabolic health have also been observed.^53^ For example, a 2021 meta-analysis of ambient air pollution exposure and blood pressure in children and adolescents concluded that short-term and long-term exposures to air pollution, specifically PM_2.5_, may increase blood pressure, but the authors stated that conclusions could not be drawn for O_3_ due to the limited number of studies.^54^ Our results suggest that investment in additional studies of O_3_ is warranted. Additionally, future work should evaluate whether more recent exposures or long-term, cumulative exposures are more relevant to health outcomes.

Although the magnitudes of the associations between air pollution exposure and health outcomes estimated here were small, air pollution exposure is ubiquitous, and elevated odd ratios applied to an entire population translate into substantial impacts on public health. Moreover, air pollution exposure is modifiable, and the public health risk it poses can be mitigated through regulation, e.g., more stringent ambient air quality standards. To this point, Brauer et al (2021) recently argued that because air pollution exposure is pervasive, reducing pollutant concentrations presents a powerful and critical opportunity to *equitably* reduce cardiovascular disease burden.^13^ The European Society of Cardiology also recognized air pollution exposure as a major modifiable risk factor relevant to the prevention of cardiovascular disease and emphasized the need for research regarding the role of air pollution in relation to early markers of cardiovascular health such as hypertension.^55^

Multiple cohort studies in the US have explored associations between air pollution exposures and cardiovascular and/or cardiometabolic health outcomes. To date, most have focused on middle aged and older adults with minimum ages of 45–50 years and/or are non-representative or small, geographically localized samples. Examples include the Multi-Ethnic Study of Atherosclerosis (MESA),^5,56^ Atherosclerosis Risk in Communities (ARIC),^57^ the Framingham Heart Study,^58^ the Jackson Heart Study,^59^ the Women’s Health Initiative (WHI),^60^ and the Study of Women’s Health Across the Nation (SWAN),^61^ among others. In these cohorts, associations have been observed between exposures to PM_2.5_ and/or O_3_ (e.g., ≥1 year) and: fatal and non-fatal cardiovascular events;^60^ diabetes;^62^ impaired renal function;^63^ systemic inflammation;^58^ and arterial injury and subclinical markers of arterial disease that are predictive of coronary heart disease and stroke in individuals without cardiovascular disease.^7^

The present study of the Add Health cohort is one of few that examines long-term air pollution exposures and cardiometabolic health outcomes in a national cohort of young adults from communities across the US. It adds to a growing body of literature on air pollution exposure and cardiometabolic health in children and young adults. For example, a study of 158 individuals in Southern California aged 17–22 years, found that one-year NO_2_ exposure was associated with higher fasting serum lipid measures (e.g., total cholesterol) and one-month O_3_ exposure was associated with higher triglyceride levels and lower HDL levels.^64^ Results of another study in Southern California that examined 173 young adults aged 18– 23 years suggested that long-term (e.g., ≥1 year) air pollution exposure may contribute to the metabolic dysfunction in youth through lipolysis and altered fatty acid metabolism.^65^ A third study from California observed that higher annual average PM_2.5_ exposure over 3 years of follow up was associated with more rapid declines in insulin sensitivity over time and lower insulin sensitivity, independent of adiposity, in overweight and obese children aged 8–15 years at enrollment.^66^ A longitudinal study of approximately 3,300 children in multiple Southern California communities enrolled at 9-10 years of age and followed until age 18 found that traffic density was associated with BMI at age 18, particularly in females.^67^ Although we did not observe associations between PM_2.5_, O_3_, and cholesterol specifically, our results add to the existing literature indicating long term air pollution exposure is associated with multiple measures of cardiometabolic health, even in young adults.

In addition to associations between air pollution exposure and health, we also observed differences in PM_2.5_ and O_3_ exposure by race/ethnicity. Non-Hispanic Black participants (and non-Hispanic White participants) had the highest O_3_ exposure, while Other race group participants had the lowest O_3_ exposure. Non-Hispanic Black participants also had the highest PM_2.5_ exposure, while non-Hispanic White participants had the lowest PM_2.5_ exposures. These findings are consistent with literature examining racial/ethnic disparities in air pollution exposure, which have generally found that non-Hispanic Blacks have higher exposures to air pollutants, especially PM_2.5_.^68-71^ This disparity may reflect, among other things, documented patterns of siting hazardous waste sites, polluting industrial facilities, and other undesirable activities or contaminated land-use types disproportionately in communities with lower socioeconomic status and a higher proportion of minoritized racial/ethnic groups.^71,72^ For this reason, it has been argued that reducing air pollution exposure is one potential pathway for reducing racial/ethnic disparities in cardiometabolic health.^13^

This study has several limitations. Add Health collects current residential address at each wave but does not collect residential histories. Thus, we do not have between-wave residential address information. Average air pollution exposures were estimated assuming that individuals resided at their Wave III residential address between 2002–05, and Wave IV residential address between 2006–07. Additionally, the FAQSD data attached to the Add Health data and used to estimate air pollution exposures was only available at the census tract level and are not available prior to 2002. Air pollution estimates continue to evolve with increasingly higher resolution and made publicly available. Future studies of high-resolution estimates, including data prior to 2002, are needed in Add Health and other cohorts.

However, this study has important strengths. This is the first study of air pollution exposures and health outcomes in the Add Health cohort. This cohort represents a large national sample of young people that includes individuals from across the US, in both urban and rural environments, and is a longstanding cohort with rich individual- and contextual-level longitudinal data. Although associations between air pollution exposure and cardiovascular and cardiometabolic health have been evaluated in other study cohorts, prior results were derived from overwhelmingly middle aged and older cohorts and/or were based on small or local samples. Examining health outcomes earlier in the life course may provide more insight into the relevance of environmental exposures in developing early markers of cardiometabolic health (or disease) and thus more opportunities for early, targeted, and/or preventative or mitigative intervention.

This study adds to the body of evidence that exposures of air pollutants may be key risk factors for cardiovascular disease, including hypertension, high BMI, diabetes, inflammation, and metabolic syndrome in early life.^26,53^ Biological mechanisms through which air pollution influences development of cardiovascular and cardiometabolic disease, or risk factors for disease have been postulated, including increased systemic inflammation,^73,74^ platelet activation in the bloodstream,^75,76^ autonomic nervous system changes,^77,78^ alterations in the vascular cell type composition,^74^ and alterations in the gut microbiome,^79^ but overall remain poorly understood. Future research would benefit from bringing together longitudinal studies representing a broad lifespan (childhood, adolescent, and adulthood) with repeated biological assessments, precisely measured environmental exposures capturing critical development periods, and clinical follow-up to better understand if and how long-term air pollution exposures shape cardiovascular and cardiometabolic disease risks across the life course.

## Data Availability

All air pollution exposure data are publicly available online: https://www.epa.gov/hesc/rsig-related-downloadable-data-files.
All health-related data is available through the Add Health Survey, which has both restricted and public use data sets. Data used here was part of the restricted use data set, access to which can be obtained through applying at the Data Portal: https://addhealth.cpc.unc.edu/data/#restricted-use. Written informed consent was obtained at all waves and the study and study protocols were approved by the Institutional Review Board of the University of North Carolina at Chapel Hill.

