## Supplemental Material for "Long-term air pollution exposure and markers of cardiometabolic health in the National Longitudinal Study of Adolescent to Adult Health (Add Health) Study"

Author affiliations: [1] Global Health Institute, School of Medicine, Duke University, Durham, NC, USA; [2] GenOmics, Bioinformatics, and Translational Research Center, RTI International, Research Triangle Park, NC, USA; [3] Fellow Program, RTI International, Research Triangle Park, NC, USA; [4]Carolina Population Center, University of North Carolina at Chapel Hill, Chapel Hill, NC, USA; [5]Department of Sociology, University of North Carolina at Chapel Hill, Chapel Hill, NC, USA.

**Ambient air pollution data**

Input data to the downscaler model that generates the FAQSD data used here includes: (1) air pollutant monitoring data obtained from the US EPA National Ambient Monitoring System/State and Local Ambient Monitoring System network (including stations with some data missingness); and (2) simulated air pollutant data in the form of gridded CMAQ numerical output. Output from CMAQ used as input to the downscaler include $O_3$ and $PM_{2.5}$ concentrations simulated at 12 × 12 km grid cells. The CMAQ model is a deterministic regional air quality model using nonlinear partial differential equations to mathematically approximate underlying physical and chemical processes occurring in the atmosphere. Utilizing output from a meteorological model and an emissions inventory, CMAQ simulates chemical and physical atmospheric processes to model pollutant transformation, transport, and fate, producing estimates of pollutant concentrations and deposition fluxes at different horizontal resolutions and atmospheric layers.[80] The meteorological inputs to the CMAQ simulations used in the downscaler statistical model are from the Mesoscale Model. The emissions inventory is based on the National Emissions Inventory.[81] Archived daily FAQSD data are publicly available from the US EPA, and additional details on the modeling approach, validation, and results are available elsewhere.[42,43]

**SM Figure S1. Sample size restrictions flow chart for sensitivity analysis**

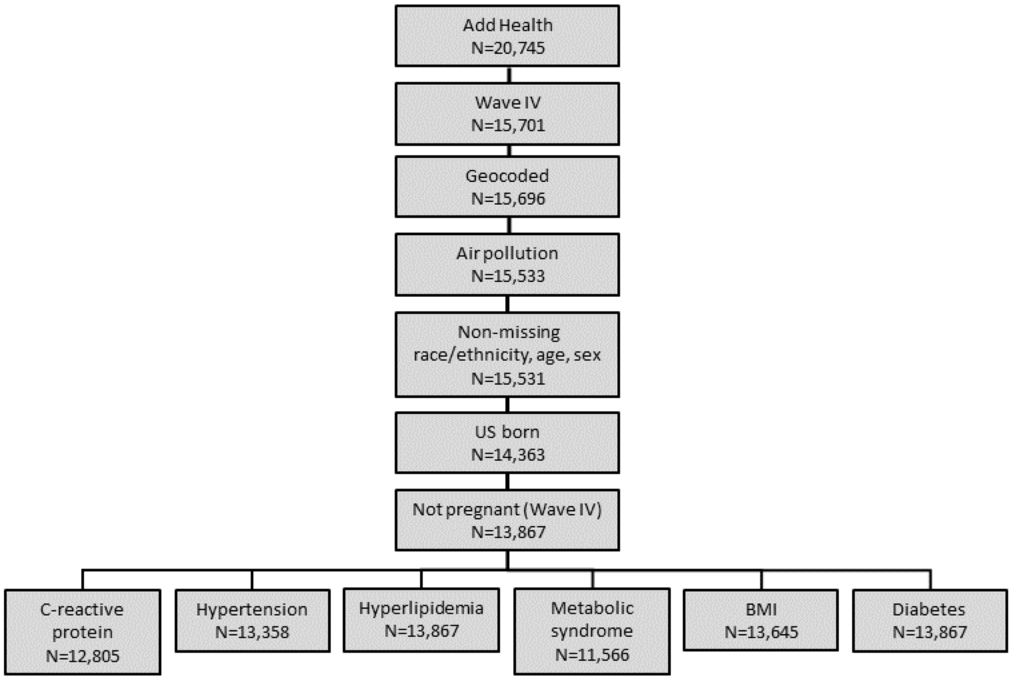

| **SM Table S1. Distributions of study participants' characteristics of Wave IV participants (n=13,867)[a]** | |
|---|---|
| **Variable** | **Mean value (SD)** |
| Age at exam, Wave IV, years | 28.4 (1.78) |
| $O_3$ exposure (ppb), 2006-2007 | 39.1 (4.38) |
| $PM_{2.5}$ exposure (μg/$m^3$), 2006-2007 | 12.2 (2.38) |
| | **N (%)** |
| Women, % | 7,166 (51.7) |
| Race/ethnicity | |
| Non-Hispanic Black | 3,290 (23.7) |
| Non-Hispanic White | 8,339 (60.1) |
| Hispanic | 1,588 (11.5) |
| Other | 650 (4.69) |
| Health outcomes | |
| Hypertension | 3,503 (26.2) |
| Hyperlipidemia | 1,140 (8.22) |
| Obese (BMI $\geq$ 30) | 5,108 (37.4) |
| Diabetes | 998 (7.20) |
| High inflammation | 4,963 (38.8) |
| Metabolic syndrome | 2,812 (20.3) |

[a] Summary statistics are presented for the analytic sample size of Wave IV only participants (n=13,867).

| **SM Table S2. Wave IV health outcomes and air pollution exposure (2006-2007) in adjusted GEE logistic regression models [a]** | | | | |
|---|---|---|---|---|
| | **Ozone** | | **$PM_{2.5}$** | |
| Health outcome (n) | OR (95% CI) | p-value | OR (95% CI) | p-value |
| Hypertension (n=13,358) | 1.003 (0.992, 1.013) | 0.628 | 1.020 (1.000, 1.041) | 0.0518 |
| Hyperlipidemia (n=13,867) | 1.005 (0.990, 1.020) | 0.515 | 1.016 (0.989, 1.044) | 0.246 |
| Obese (n=13,645) | **1.014 (1.003, 1.025)** | **0.00993** | 1.001 (0.980, 1.023) | 0.920 |
| Diabetes (n=13,867) | **1.024 (1.004, 1.045)** | **0.0198** | 0.999 (0.965, 1.034) | 0.936 |
| Inflammation (n=12,805) | **1.012 (1.003, 1.022)** | **0.0102** | 0.996 (0.979, 1.014) | 0.677 |
| Metabolic syndrome (n=11,566) | **1.013 (1.001, 1.026)** | **0.0409** | 1.004 (0.979, 1.030) | 0.743 |

[a] GEEs were fit as logistic regression models with a binary health outcome, PSUSCID as the cluster ID variable, and an exchangeable correlation matrix. Models adjusted for pollutant concentration, age, sex, and race/ethnicity (non-Hispanic Black, non-Hispanic White, Hispanic, and Other). Note sample sizes vary some from largest N shown here.
